# Sociodemographic Predictors of Consent: A Protocol and Statistical Analysis Plan for a Nested Observational Study of Canadian Sites in the REVISE Trial

**DOI:** 10.64898/2026.07.06.26357216

**Authors:** Nicholas Bauer, Alexandra Binnie, Vinayak Lad, Rosa Myrna Marticorena, Jennifer Tsang, Nicole Zytaruk, Diane Heels Ansdell, Deborah Cook, the Canadian Critical Care Trials Group

## Abstract

**Background:** In Canada, there is a lack of data relating sociodemographic characteristics to the likelihood of consent and clinical trial participation.

**Objective:** The overall objective of this study is to examine the association of hospital-level sociodemographic variables with *a priori* informed consent rates for participation in the REVISE trial.

**Design:** This study is a retrospective observational analysis of Canadian sites participating in the international REVISE trial.

**Methods:** Sociodemographic characteristics for 42 hospitals participating in the REVISE trial will be supplemented by national data from the 2021 Canadian Census of Population Profile at the census tract level corresponding to the hospital’s location. Hospital level information for Ontario sites will be derived from the Institute for Clinical Evaluate Sciences (ICES) database. Site clustering will be performed using latent class analysis, a flexible clustering technique that identifies meaningful subgroups based on sociodemographic variables purposively selected from data available through the Statistics Canada 2021 census profile, ICES, and hospital-reported data. Clustering analysis will be performed for all Ontario hospitals with available ICES data, followed by a separate analysis for all Canadian REVISE sites using Statistics Canada data. Concordance in the clustering of REVISE sites will be examined by comparing the assignment of hospitals to the latent classes separately identified using ICES and Statistics Canada data. If there is a high degree of agreement between the two datasets, sociodemographic predictors will be analyzed using the clusters identified through ICES for Ontario sites with the concordant classes based on Statistics Canada data for Canadian sites outsite Ontario. If there is disagreement in cluster assignment between the two datasets, separate analyses of sociodemographic factors will be conducted for Ontario sites using ICES data and for all Canadian sites using the 2021 Census Profile. Multivariate linear regression models will be used to analyze the association between hospital-level characteristics and the likelihood of *a priori* and deferred consent.

**Results:** Results of this study will generate information about the relationship between informed consent to participate in a low-risk critical care clinical trial using different consent models, and socioeconomic patient characteristics at the hospital site level (e.g., educational attainment, knowledge of official languages, citizenship rates, family income, poverty, rurality and immigration patterns).

**Conclusions:** This study will fill an evidence gap by generating information on the relationship between sociodemographic variables and the likelihood of informed consent to participate in a critical care clinical trial in Canada.

## BACKGROUND

Retrospective analyses have identified lower clinical trial participation amongst patients who are structurally vulnerable, including those with socioeconomic disadvantage, racialized identities, lower health literacy and educational attainment, lower proficiency in official languages, and recent immigration status.^1-9^ In intensive care unit (ICU) settings, these same sociodemographic factors have been associated with higher rates of ICU admission, greater disease severity, higher ICU mortality, longer length of stay, and worse outcomes following critical illness.^10-16^ Thus, patient populations at higher risk of poor outcomes may be systematically underrepresented in clinical investigations, exacerbating health inequity and introducing selection bias in research.

A key step in the recruitment of participants into research projects is obtaining informed consent. Informed consent typically occurs *a priori*, before any research interventions. It involves a process of consultation between either the patient or their substitute decision maker (SDM) and research staff. The purpose of the informed consent process is to share the objective of the study, review trial procedures, and discuss the potential risks and benefits of participation. Studies from the United States have shown that patients with lower socioeconomic status and racialized identity are less likely to participate in clinical trials.^17-19^ In Canada, there is a lack of data relating sociodemographic characteristics to the likelihood of consent and clinical trial participation.

The REVISE trial (<u>R</u>e-<u>Ev</u>aluating the Inhibition of <u>S</u>tress <u>E</u>rosions) was an international, multicentre, randomized, placebo-controlled study to determine, among invasively mechanically ventilated patients, the effect of pantoprazole versus placebo on the primary efficacy outcome of clinically important upper gastrointestinal bleeding, and the primary safety outcome of 90-day mortality.^20^ The trial found that pantoprazole significantly reduced both clinically important and patient-important gastrointestinal bleeding, with no effect on 90-day mortality. Of the 4,821 patients enrolled in REVISE, 3,265 were enrolled in Canada across 42 sites, including 30 academic hospitals and 12 community hospitals. Sociodemographic variables were not collected for individual patients enrolled in REVISE, however hospital-level data can provide some insight into the characteristics of patients enrolled across participating sites and the relationship between socioeconomic variables and the likelihood of informed consent to participate in this trial.

### Objectives

The overall aim of this study is to examine the association of hospital-level sociodemographic variables with *a priori* informed consent rates for participation in the REVISE trial, with 5 specific objectives.

- <u>Objective 1</u>: Identify hospital-level sociodemographic characteristics using data obtained from Statistics Canada (for all Canadian sites) and the Ontario Ministry of Health’s ICES AHRQ Program (for Ontario hospitals).
- <u>Objective 2</u>: Perform clustering analysis to identify hospitals with similar sociodemographic and institutional characteristics
- <u>Objective 3</u>: Assess concordance between latent classes identified independently using ICES and Statistics Canada data.
- <u>Objective 4</u>: Compare *a priori* informed consent rates between hospital clusters using REVISE site consent data
- <u>Objective 5</u>: Perform logistic regression analysis to identify hospital-level characteristics which are associated with REVISE *a priori* consent rates

### Hypothesis

We hypothesize that lower *a priori* consent rates will be associated with site-level sociodemographic factors, including: lower educational attainment, lower knowledge of official languages, lower citizenship rates, lower median after-tax family income, higher burdens of poverty, higher rurality and higher proportion of recent immigrants (last 10 years).

## Methods

### Study Population

This study is a retrospective observational analysis of Canadian sites participating in the international REVISE trial.^20^ REVISE recruited eligible adults (over 18 years of age) undergoing invasive mechanical ventilation in participating ICUs. According to approvals by local research ethics boards, eligible participants were enrolled into the study by *a priori* informed consent, deferred consent, or using an opt-out approach in one international site. Study participants were randomized in a 1:1 ratio to receive intravenous pantoprazole or placebo. Pantoprazole or placebo were administered in a blinded manner for 90 days or until mechanical ventilation was discontinued, or a prespecified indication or contraindication for proton pump inhibitors emerged, or death, whichever outcome occurred first.

### Source Data

Sociodemographic characteristics for each hospital were derived from a hospital-reported survey completed by REVISE Research Coordinators. This information will be supplemented by national data from the 2021 Canadian Census of Population Profile at the census tract level corresponding to the hospital’s location.^21,22^ Hospital level information for sites in Ontario will be derived from the Institute for Clinical Evaluate Sciences (ICES) databases, accessed through the Ontario Ministry of Health’s Applied Health Research Question (AHRQ) Program.^23,24^

### Statistical Analysis

The sample size for this sub-study is fixed, based on 42 participating Canadian hospitals. Site data will be presented using descriptive statistics with means and standard deviations or medians and interquartile range as appropriate for continuous data, and absolute counts and percentages for categorical data. Site clustering will be performed using latent class analysis (LCA), a flexible clustering technique that identifies meaningful subgroups based on sociodemographic variables purposively selected from data available through the Statistics Canada 2021 census profile, ICES, and the hospital-reported survey. Clustering analysis will be performed for all Ontario hospitals with available ICES data, followed by a separate analysis for all Canadian REVISE sites using Statistics Canada data. Concordance in the clustering of REVISE sites will be examined by comparing the assignment of hospitals to the latent classes separately identified using ICES and Statistics Canada data. If there is a high degree of agreement between the two datasets, sociodemographic predictors will be analyzed using the clusters identified through ICES for Ontario sites with the concordant classes based on Statistics Canada data for Canadian sites outsite Ontario. If there is disagreement in cluster assignment between the two datasets, separate analyses of sociodemographic factors will be conducted for Ontario sites using ICES data and for all Canadian sites using the 2021 Census Profile.

These analyses will be considered exploratory. Statistical comparisons will be performed using the χ2 test for categorical variables, Student t-test for continuous parametric variables, and Mann-Whitney rank sum test for continuous non-parametric variables. Multivariate linear regression models will be used to analyze the association between hospital-level characteristics and the likelihood of *a priori* and deferred consent. All tests will be two-tailed and an alpha of 0.05 will be considered significant. Statistical analysis will be conducted using the R computing platform.

## Ethics and Dissemination

Protocol implementation for the REVISE trial and database training aligned with the International Council for Harmonisation Guidelines for Good Clinical Practice and other locally applicable regulations. The REVISE protocol was registered on clinicaltrials.gov (NCT 03374800), was approved by Health Canada and all research ethics boards associated with participating centers. Free and informed consent was obtained for research participants in this trial. The REVISE Consent Substudy,^25^ the results of which will be the foundation of the current protocol,^26^ was approved by the Hamilton Integrated Research Ethics Board (#19124).

## Results

Results of this study will generate information about the relationship between socioeconomic patient characteristics at the hospital site level and the likelihood of informed consent to participate in a low-risk critical care clinical trial.

## Discussion

This study builds on policy work that enhances and promotes the inclusion of structurally vulnerable patients in clinical trials. Strengths of this protocol include the large number of hospital sites from across Canada, including 12 community hospitals, along with detailed socioeconomic site data from ICES and the Ministry of Health of Ontario and detailed clinical and outcome data from the REVISE trial. In addition, this pre-specified protocol and statistical analysis plan helps to ensure methodological rigour. Finally, the broad Canadian representation will enhance the generalizability of the findings to other critical care trials across the country.

We will share results with the Canadian Critical Care Trials Group. We will publish these findings through conventional academic channels, including abstracts of critical care meetings and peer-reviewed manuscripts. We will also disseminate results at professional fora with research trainees, and social media channels.

## Conclusions

This study will fill an evidence gap by generating information on the relationship between sociodemographic variables and the likelihood of informed consent to participate in a critical care clinical trial in Canada.

## Data Availability

All data produced in the present study are available upon reasonable request to the authors

